# Deciphering the Role of Complement System Genes in Pancreatic Cancer Susceptibility and Prognosis

**DOI:** 10.1101/2025.04.17.25326022

**Authors:** A Langtry, R Rabadan, L Alonso, I Filip, S Sabroso, A Moreno-Oya, R Lawlor, A Carrato, R Alvarez-Gallego, M Iglesias, X Molero, MJ Löhr, CW Michalski, J Perea, M O’Rorke, VM Barberà, A Tardón, A Farré, L Muñoz-Bellvís, T Crnogorac-Jurcevic, E Domínguez-Muñoz, T Gress, W Greenhalf, L Sharp, J Balsells, E Costello, J Kleeff, B Kong, J Mora, D O’Driscoll, A Scarpa, W Ye, FX Real, E López de Maturana, N Malats, PanGenEU Investigators

**Affiliations:** Genetic and Molecular Epidemiology Group, Spanish National Cancer Research Center (CNIO), and CIBERONC, Madrid, Spain; Department of Systems Biology, Columbia University, New York, NY, USA; ARC-Net Center for Applied Research on Cancer, Department of Diagnostics and Public Health and Department of Engineering for Innovation Medicine, University of Verona, Verona, Italy; Department of Oncology, Ramón y Cajal University Hospital, IRYCIS, Alcala University, Madrid, and CIBERONC, Spain; HM CIOCC Madrid (Centro Integral Oncológico Clara Campal). Hospital Universitario HM Sanchinarro. Instituto de Investigación Sanitaria HM Hospitales. Facultad HM de Ciencias de la Salud de la Universidad Camilo José Cela.; Department of Pathology, Hospital del Mar—Parc de Salut Mar, Barcelona, and CIBERONC, Spain; Department of Gastroenterology, Hospital Vall Hebron, Barcelona; and Hospitals Universitaris Arnau de Vilanova i Santa Maria, IRBLleida, UdL, 25198, Lleida; and CIBEREHD, Madrid, Spain; Gastrocentrum, Karolinska Institutet and University Hospital, Stockholm, Sweden; Department of Surgery, Technical University of Munich, Munich; and Department of General, Visceral and Transplantation Surgery, Medical Faculty Heidelberg, University Heidelberg, Germany; Department of Surgery, Hospital 12 de Octubre, Madrid; and Institute of Biomedical Research of Salamanca, Salamanca, Spain; Centre for Public Health, Belfast, Queen’s University Belfast, UK; and College of Public Health, The University of Iowa, Iowa City, IA, US; Molecular Genetics Laboratory, General University Hospital of Elche, Spain; Department of Medicine, Instituto Universitario de Oncología del Principado de Asturias (IUOPA), Instituto de Investigación Sanitaria del Principado de Asturias (ISPA), Oviedo; and CIBERESP, Madrid, Spain; Departments of Gastroenterology and Clinical Biochemistry, Hospital de la Santa Creu i Sant Pau, Barcelona, Spain; Department of Surgery, Hospital Universitario de Salamanca – IBSAL, Universidad de Salamanca, Salamanca; and CIBERONC, Madrid, Spain; Barts Cancer Institute, Centre for Cancer Biomarkers and Biotherapeutics, Queen Mary University of London, London, UK; Department of Gastroenterology, University Clinical Hospital of Santiago de Compostela, Spain; Department of Gastroenterology, University Hospital of Giessen and Marburg, Marburg, Germany; Department of Molecular and Clinical Cancer Medicine, University of Liverpool, Liverpool, UK; National Cancer Registry Ireland and HRB Clinical Research Facility, University College Cork, Cork, Ireland; and Newcastle University, Institute of Health & Society, Newcastle, UK; Department of Gastroenterology, Hospital Vall Hebron, Barcelona, Spain; Department of Surgery, Technical University of Munich, Munich; and Department of Visceral, Vascular and Endocrine Surgery, Martin-Luther-University Wittenber-Halle (Saale), Germany; National Cancer Registry Ireland and HRB Clinical Research Facility, University College Cork, Cork, Ireland; Department of Medical Epidemiology and Biostatistics, Karolinska Institutet, Stokholm, Sweden; Epithelial Carcinogenesis Group, Spanish National Cancer Research Center (CNIO) and CIBERONC, Madrid; and Pompeu Fabra University, Barcelona, Spain; In Annex 1

## Abstract

**Background & Aims:** Pancreatic ductal adenocarcinoma (PDAC) genetic susceptibility is partially identified. The complement system (CS) influences carcinogenesis and participates in immunological defense and homeostasis; however, its role in PDAC genetic susceptibility and prognosis is underexplored.

**Methods:** The association of SNPs within 111 CS-related genes with PDAC risk was assessed in the PanGenEU study and validated in the UKBiobank. We investigated the association between the CS-related gene variation and PDAC risk, followed by an in-depth functional *in-silico* study using TCGA and ICGC data. We assessed whether CS-related genes were associated with prognosis at germline and somatic levels. We investigated the immune infiltration of PDAC tumors according to their transcriptomic profile.

**Results:** Genetic variation in *FCN1* and *PLAT* was significantly associated with PDAC risk. PDAC patients with elevated expression of *IGHG3*, *IGKC*, *IGHM*, *F2R*, *F2RL2*, *CFI*, *A2M*, and *C4A* displayed improved survival and higher infiltration of CD8^+^, B cells, and Th1 cells. Individuals with high expression levels of *FGA*, *SERPINE1*, *FGG*, and *F3* had poorer survival, higher infiltration of Tregs, and lower infiltration of CD8+ cells.

**Conclusions:** Results from this study suggest that CS-related genes play a role in PDAC genetic susceptibility and survival through specific immune cell infiltration.

---

Pancreatic cancer remains one of the most lethal cancers which is predicted to become the second leading cause of cancer-related deaths by 2030^1,2^. Pancreatic ductal adenocarcinoma (PDAC) accounts for 95% of pancreatic cancer cases, and patients with PDAC display a dismal 5-year survival rate of 5-10%^3,4^. Unfortunately, this bleak scenario will not change in the future unless multiple actions are taken to control PDAC, including the identification of PDAC high-risk populations to improve primary and secondary prevention interventions. To this end, a better understanding of the role of genetic and non-genetic factors involved in pancreas carcinogenesis is central.

The immune system plays a critical role in detecting and eliminating cancer cells; however, it can also promote tumor growth and progression through various mechanisms^5,6^. Among the immune-related pathways, multiple pieces of evidence support the role of the Major Histocompatibility Complex (MHC) region in PDAC risk. While most studies have focused on the MHC class I and class II regions, the MHC III region has barely been explored. This region is responsible for inflammatory processes and the CS response, a multistep cascade operating as a key component of the innate immune response. Its activation triggers the formation of the cell-killing membrane attack complex (MAC), which disrupts the cell membrane and ultimately leads to the lysis of target cells. This process is orchestrated through three distinct activation pathways: classical, lectin, and alternative pathways^7–9^. Further to this role, the complement cascade interacts closely with the clotting system. There is strong evidence that several proteases of the coagulation system can activate C3 and C5, indirectly leading to the activation of the CS^10^. Notably, the CS can be activated by cancer cells and their microenvironment, producing inflammatory and immunomodulatory molecules that can promote tumor growth and suppress immune surveillance^5,11,12^. Genetic variation within CS-related genes, such as C3, C5^13^, or CD35^14^, have been associated with the prognosis of different cancer types, including PDAC^15^. In fact, some CS genes have been suggested as biomarkers for PDAC prognosis and treatment response^16^. However, the role of the CS in cancer risk and prognosis, particularly in the context of PDAC, is still not well understood. This is particularly relevant given the fact that PDAC risk also associates with asthma, allergies, autoimmune diseases, and other inflammatory conditions^17,18^.

In this study, we report on the association between SNPs within the CS-related genes and the genetic susceptibility to and prognosis of PDAC. Considering that the impact of a single genetic variant might have a small effect on a complex disease like PDAC, we conducted a comprehensive gene-based association analysis by aggregating all variants within each gene to provide a broader perspective on the overall gene-level effects. To gain deeper insights into the identified susceptibility loci in PDAC, we have also integrated the information on the association of SNPs with other omics data such as expression and splicing quantitative trait loci (eQTL and sQTL, respectively) in normal pancreas using colocalization and performing functional *in silico* analysis. Overall, our findings suggest that several CS-related genes play a significant role in the genetic susceptibility to PDAC. Finally, we demonstrate that the expression of different CS-related genes not only influence PDAC survival but it also correlates with immune cell infiltration patterns in PDAC. We also provide a signature of genes the expression of which is associated with improved PDAC prognosis. By utilizing large-scale studies and state-of-the-art analytical methodologies, our study contributes to a deeper understanding of the role of CS-related genes in pancreas carcinogenesis and progression shedding light on the potential implications for disease susceptibility and prognosis.

## MATERIAL and METHODS

### Ethics Statement

IRB ethical approval was obtained by all participating centers contributing to PanGenEU. Written informed consent was obtained from all study participants, respectively. The study was conducted in accordance with the Helsinki Declaration.

We utilized the resources of the PanGenEU case-control study, the UK Biobank, the International Cancer Genome Consortium (ICGC), Pancreatic Cancer Canada (ICGC-CA) and Pancreatic Cancer Australia (ICGC-AU) repositories (https://dcc.icgc.org/repositories), and The Cancer Genome Atlas (TCGA) repository (https://portal.gdc.cancer.gov). A description of these studies and populations can be found in **Supplementary Methods.**

### Single variant association analysis with PDAC risk

For the single variant association analysis with PDAC risk, we considered both imputed and genotyped SNPs annotated within the 111 complement-related genes preselected by Qian et al. (2019)^19^ (**Supplementary Table 2**), for both PanGenEU (197,050 SNPs) and UK Biobank (183,849 SNPs) studies. In the PanGenEU, we prioritized SNPs with a minor allele frequency (MAF) ≥5% and high imputation quality (info >0.3). To estimate the OR and 95% CI for the selected SNPs with PDAC risk in each study population, we conducted logistic regression models adjusted for the first five principal components to account for population stratification, age, gender, and the European region as covariates in the case of the PanGenEU study population. For the UK Biobank population, we adjusted the models for the aforementioned variables except for the European region, in addition to the genotype array type and center. Finally, we combined the individual study estimates in a meta-analysis assessing the summary statistics of the association analysis of 5,932 SNPs obtained in both populations using the function *rma.umi* (metafor R package, v. 4.2-0)^20^.

### Gene-based association analysis with PDAC risk

We conducted a gene-based association analysis in each population using the Sequence Kernel Association Test (SKAT-O), implemented in the R package *seqMeta* (v1.6.7) (https://github.com/DavisBrian/seqMeta). To do this, we employed the SNPs included in the single variant association analysis, followed by a filtering step based on linkage disequilibrium (LD>0.7). We adjusted the SKAT-O models for the same covariates as mentioned previously. Furthermore, to detect potential associations and gain more robust insights into the role of the complement-related genes in PDAC risk, we performed a meta-analysis of the summary statistics from the 111 complement-related genes derived from both study populations. We then assessed the association (OR and 95%CI) between the CS-related genes linked with PDAC risk in our analysis and the well-known PC risk factors, including asthma, allergies, tobacco, family history of cancer, and body mass index (BMI) in each study population using logistic regression models adjusted for the same covariates as previous models. Furthermore, we conducted a meta-analysis using a random-effects model to integrate the results from both study populations.

To understand the functional implications of the genes significantly associated with PDAC risk, we conducted an *in silico* functional analysis using various bioinformatics tools (see **Supplementary Methods**).

### Survival analysis of CS-related genes at germline and somatic levels

We examined the association between the SNPs within 111 CS-related genes and PDAC overall survival (OS) in the PanGenEU and UK Biobank populations (see **Supplementary Methods**).

Similarly, we used data on the expression levels of the 111 CS-related genes in PDAC tumors to examine their association with the overall survival of 134 PDAC patients from TCGA, classifying them as low-expression and high-expression groups based on the median. To validate these findings, we analyzed survival data from (PanGenEU (*n*=122), ICGC-CA (*n*=160), and ICGC-AU (*n*=86)) employing Cox PH models with adjustment for sex, age at diagnosis, and tumor stage in each population. Next, we meta-analyzed the findings from the four study populations, focusing on CS-related genes with a nominal *P*< 0.1 in any of them. We also created gene expression signatures based on the meta-analysis results: *Signature 1*, comprising genes whose expression was associated with improved overall survival; *Signature 2*, comprising genes whose expression was associated with a decreased OS. These signatures were created by converting gene expression levels into a score, with patients being classified into low-expression or high-expression groups based on the median expression levels of the signature. Finally, we assessed the association between these signature scores and PDAC survival in each study population and conducted a comprehensive meta-analysis combining findings from the four cohorts.

Finally, we explored whether the observed associations between CS-related genes at the individual and signature levels with survival might be partially explained by specific immune infiltration patterns within PDAC tumors (see **Supplementary Methods**).

## RESULTS

### Characteristics of the study populations

The demographic and clinical characteristics of the PanGenEU and UK Biobank study populations are displayed in **Supplementary Table 3**. PanGenEU individuals were from Spain (58.8%), Italy (22.6%), Sweden (7.9%), Germany (6.7%), UK (3.7%), and Ireland (0.2%). The average age of cases and controls was comparable within each study population, although the UK Biobank controls were younger. Moreover, a slightly lower proportion of females was observed in both cases and controls in PanGenEU (n. s.). In terms of risk factors, cases in both PanGenEU and UK Biobank populations had a higher prevalence of smoking and diabetes compared to controls. On the other hand, the prevalence of asthma and allergies was higher among controls than among cases in both studies (**Supplementary Table 3**). We observed significant differences in the proportion of stage I-II and III-IV PDAC tumors across studies, PanGenEU was the study population with the highest proportion of stage III-IV PDAC tumors and, therefore, more representative of the full spectrum of the disease. The median survival of the TCGA patients was significantly shorter than survival in the other cohorts (PanGenEU, ICGC-AU, and ICGC-CA).

### Single variant association analyses with PDAC risk

Results of the meta-analysis of the CS-related SNPs and PDAC risk are presented in **Supplementary Table 4**. A total of 653 SNPs located within 41 CS-related genes were found to be significantly associated with PDAC risk at a nominal *P*<0.05. However, after adjusting for multiple testing (Benjamini-Hochberg), none of the SNPs remained statistically significant (minimun adjusted *P*=0.09). The five SNPs with the lowest *P* value mapper to *F13A1* (rs6597196), *A2M* (rs34803501, rs12427063, and rs12426061), and *CFH* (rs10922096).

### Gene-based association analysis with PDAC risk and established PDAC risk factors

Utilizing SKAT-O, we identified two genes significantly associated with PDAC risk in the meta-analysis, ficolin-1 (*FCN1*, adjusted *P*=9.9×10^−3^) and tissue type plasminogen activator (*PLAT*, adjusted *P*=1.56×10^−2^). Furthermore, four genes showed a borderline significant association: vitronectin (*VTN*, adjusted *P*=7.64×10^−2^), the complement regulatory protein *CD46* (adjusted *P*=7.24×10^−2^), coagulation factor II receptor-like 2 (*F2RL2*, adjusted *P*=7.24×10^−2^), and alpha-2-macroglobulin (*A2M*, adjusted *P*=7.64×10^−2^) (**Table 1**). Notably, *PLAT* and *F2RL2* were also associated with asthma and *VTN* with nasal allergies (**Supplementary Table 5**).

**Table 1.**
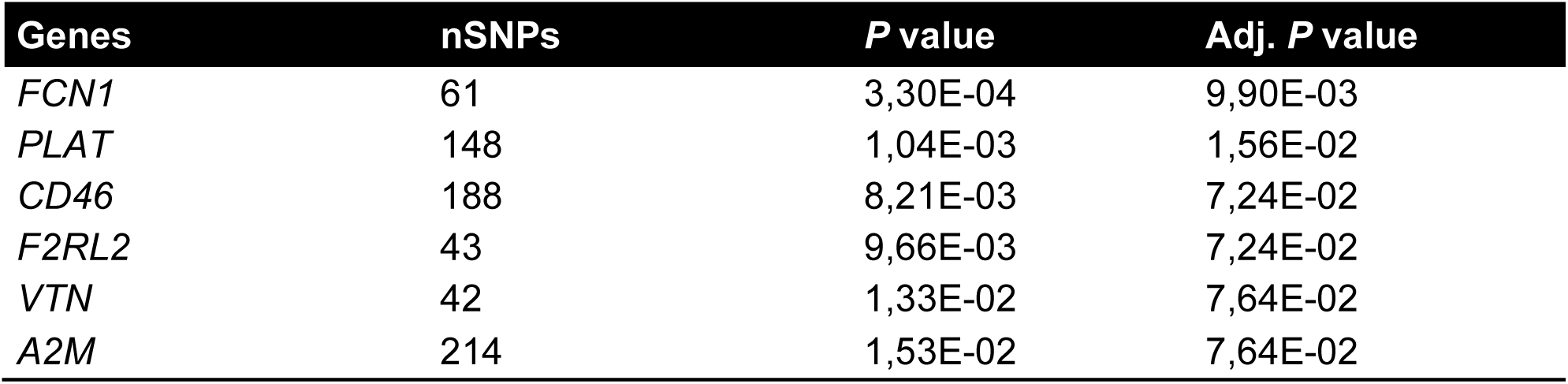
Number of SNPs (nSNPs) within each gene and *P* value of the SKAT-O association between the CS-related genes and PDAC risk.

### Functional in silico analyses

The functional *in silico* analyses provided valuable insights into the CS-related genes associated with PDAC risk. Gene set enrichment analysis conducted using FUMA GWAS revealed that pathways related to humoral immune responses, coagulation, and regulation of immune responses were enriched with the significant set of CS-related genes (**Supplementary Fig. 1**). Exploring the DisGeNET database, we found that many of the CS-related genes associated with PDAC risk had previously been associated with digestive system diseases and neoplasms, as well as with nervous diseases and mental disorders (**Supplementary Fig. 2**).

By using GTEx expression data, we discovered that within the *VTN* gene, eight out of the eleven SNPs associated with an increased risk of PDAC (nominal *P*<0.05) were also eQTLs associated with a higher expression of *VTN* in the normal pancreas: rs2227726, rs2227721, rs2227720, rs2227718, rs111502247, rs2071378, rs2071377, and rs113635549 (max. PDAC OR=1.17, *P*=4.58×10^−2^; max. eQTL OR=2.34, *P*=3.66×10^−11^) (**Supplementary Fig. 3a**). Additionally, a single SNP within *CD46* was identified as a sQTL, meaning that it is also associated with differential splicing (*CD46*-rs2796278, PDAC OR=1.08, *P*=5.79×10^−2^, sQTL OR=0.51, *P*=5.73×10^−16^) (**Supplementary Fig. 3b**). Results of colocalization analyses (**Supplementary Table 6**) suggest that there is a moderate likelihood that the causal variants associated with a higher risk of PDAC (rs2111023, rs12427063, and rs12426061) are also causal of differential expression of the *A2M* gene. The posterior probability for colocalization of PDAC-associated SNPs and eQTLs corresponding to the *A2M* gene was 46.3%. For *VTN*, the posterior probability that the SNPs are causal for both PDAC risk and expression was much lower (13.5%). For *PLAT*, *FCN1*, *CD46*, and *F2RL2* the posterior probabilities for sharing a causal variant were lower than 0.1, as well as for all the genes in the colocalization analysis of PDAC and sQTLs (**Supplementary Table 6**).

### Prognostic associations of CS-related genes in PDAC patients

We first assessed the association between the CS-related SNPs and genes with PDAC survival in both PanGenEU and UK Biobank populations independently. To further assess their association with survival, we conducted a meta-analysis incorporating the findings from each population. This identified 583 SNPs with a nominal *P* value significantly associated with PDAC survival. However, after adjusting for multiple comparisons, these associations did not retain statistical significance (**Supplementary Table 7**). Additionally, through our SKAT-O model, we also observed that three CS-related genes (*C8B*, *F2,* and *KLKB1*) were significantly associated with PDAC survival at a nominal *P* value (*P* <0.05, **Supplementary Table 8**).

### Prognostic CS-based gene signatures for patient stratification

Initially, we tested the association between the expression of CS-related genes and PDAC OS at the individual study level (PanGenEU, TCGA, ICGC-CA, and ICGC-AU). We then performed a meta-analysis combining the results from each study. Out of 17 CS-related genes associated with PDAC OS at nominal *P*<0.05, 12 remained significant after correcting for multiple testing: Ig heavy chain gamma 3 (*IGHG3*), Ig heavy chain mu (*IGHM*), Ig kappa chain (*IGKC*), coagulation factor II thrombin receptor (*F2R*), *F2RL2*, complement factor I (*CFI*), alpha-2 macroglobulin (*A2M*), complement component 4A (*C4A*), serpin family E member 1 (*SERPINE1*), fibrinogen alpha chain (*FGA*), fibrinogen gamma chain (*FGG*), coagulation factor III, tissue factor (*F3*). Results showed that an elevated expression of *IGHG3*, *IGHM*, *IGKC, F2R*, *F2RL2*, *CFI, A2M,* and *C4A* was significantly associated with better OS (**Fig. 1**), whereas higher expression of *SERPINE1, FGA*, *FGG*, and *F3* significantly associated with poorer OS (**Fig. 2**).

**Fig. 1.**
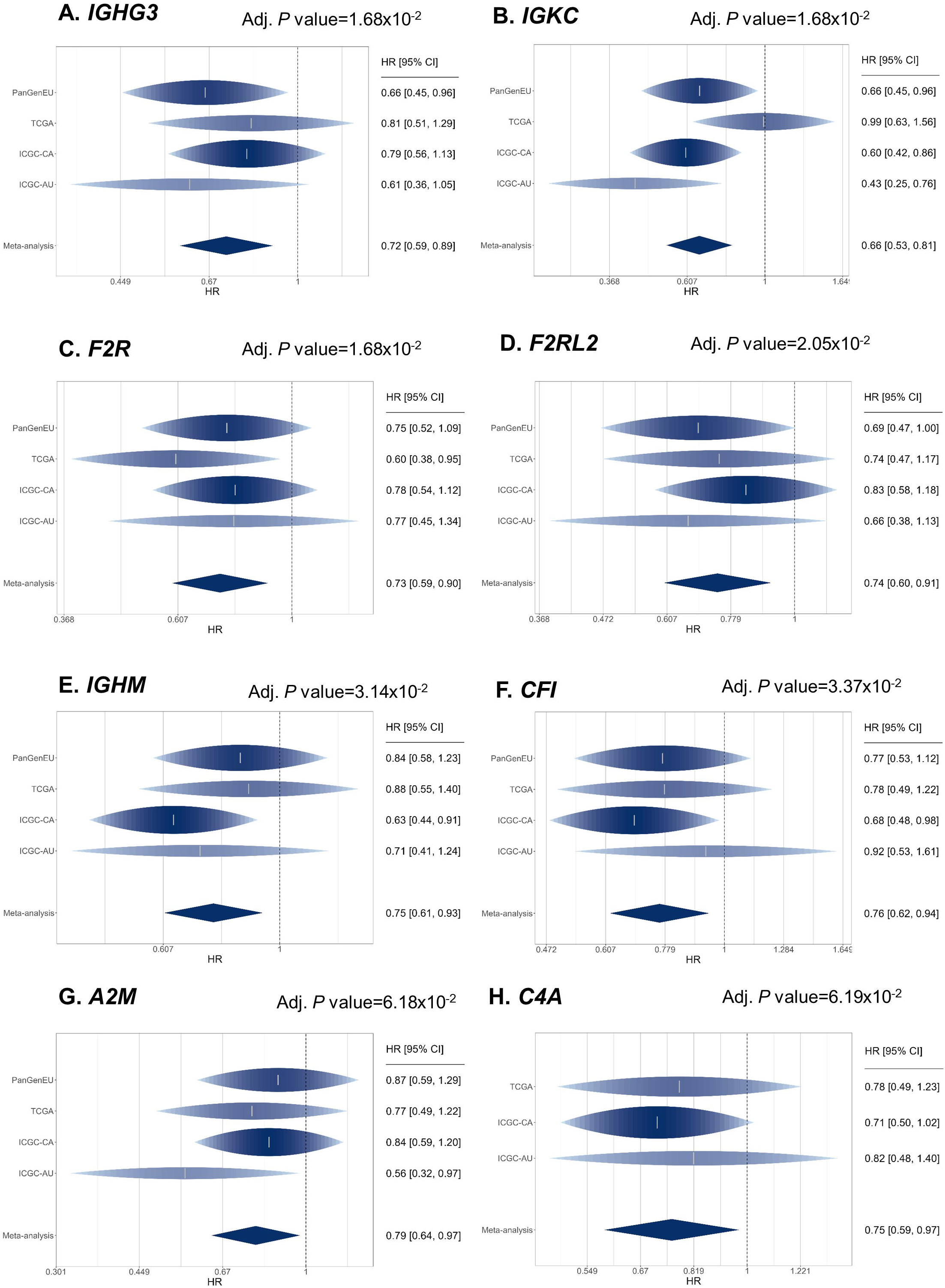
Forest plots from the meta-analysis of the CS-related genes associated with improved overall survival when considering PanGenEU, TCGA, ICGC-CA, and ICGC-AU (Figs. 1a-1g), and when including data from the TCGA, ICGC-CA, and ICGC-AU (Fig. 1h). Hazard Ratio (HR) and 95% confidence interval (CI) estimated with Cox proportional-hazards models under a dominance MoI.

**Fig. 2.**
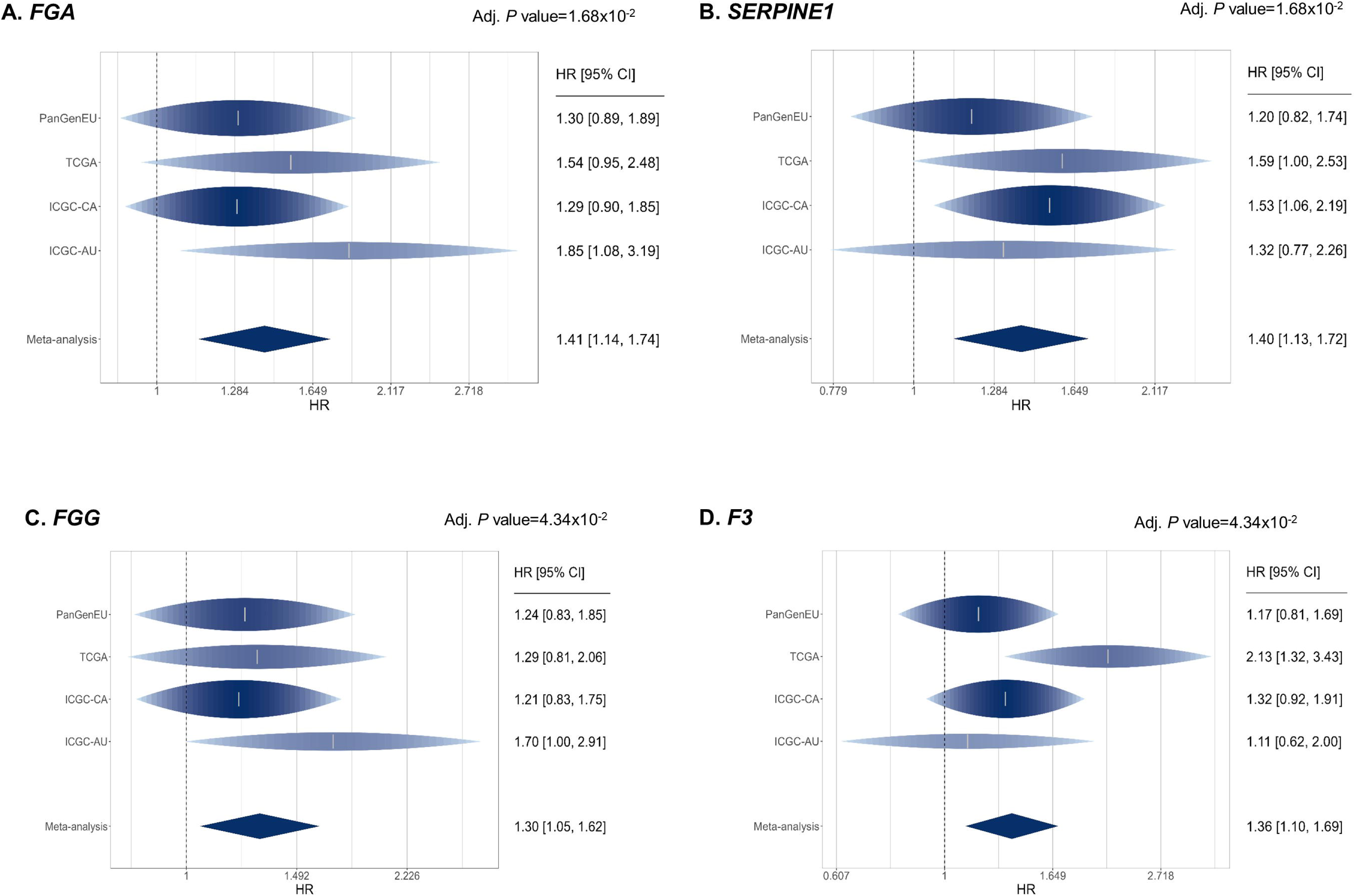
Forest plots from the meta-analysis of the CS-related genes associated with poorer overall survival when considering PanGenEU, TCGA, ICGC-CA, and ICGC-AU. Hazard ratio (HR) and 95% confidence interval (CI) were estimated with Cox proportional-hazards regression models under a dominance MoI.

We also investigated the association of two gene expression signatures comprising genes whose expression was associated with better (*Signature 1*: *IGHG3*, *IGKC*, *IGHM*, *F2R*, *F2RL2*, *CFI*, and *A2M)* or worse (*Signature 2*: *FGA*, *SERPINE1*, *FGG*, *F3*) OS in the PanGenEU, TCGA, ICGC-CA, and ICGC-AU cohorts, followed by a meta-analysis. Patients with high expression levels of *Signature 1* genes experienced a 32% reduction in mortality risk compared to those with low expression levels (HR=0.68, *P*=4.0×10^−4^, **Fig. 3a**). Conversely, patients with high expression levels of *Signature 2* genes displayed a less favorable OS compared to their low expression counterparts, although this association was not statistically significant (HR=1.16, *P*=2.01×10^−1^, **Fig. 3b**).

**Fig. 3.**
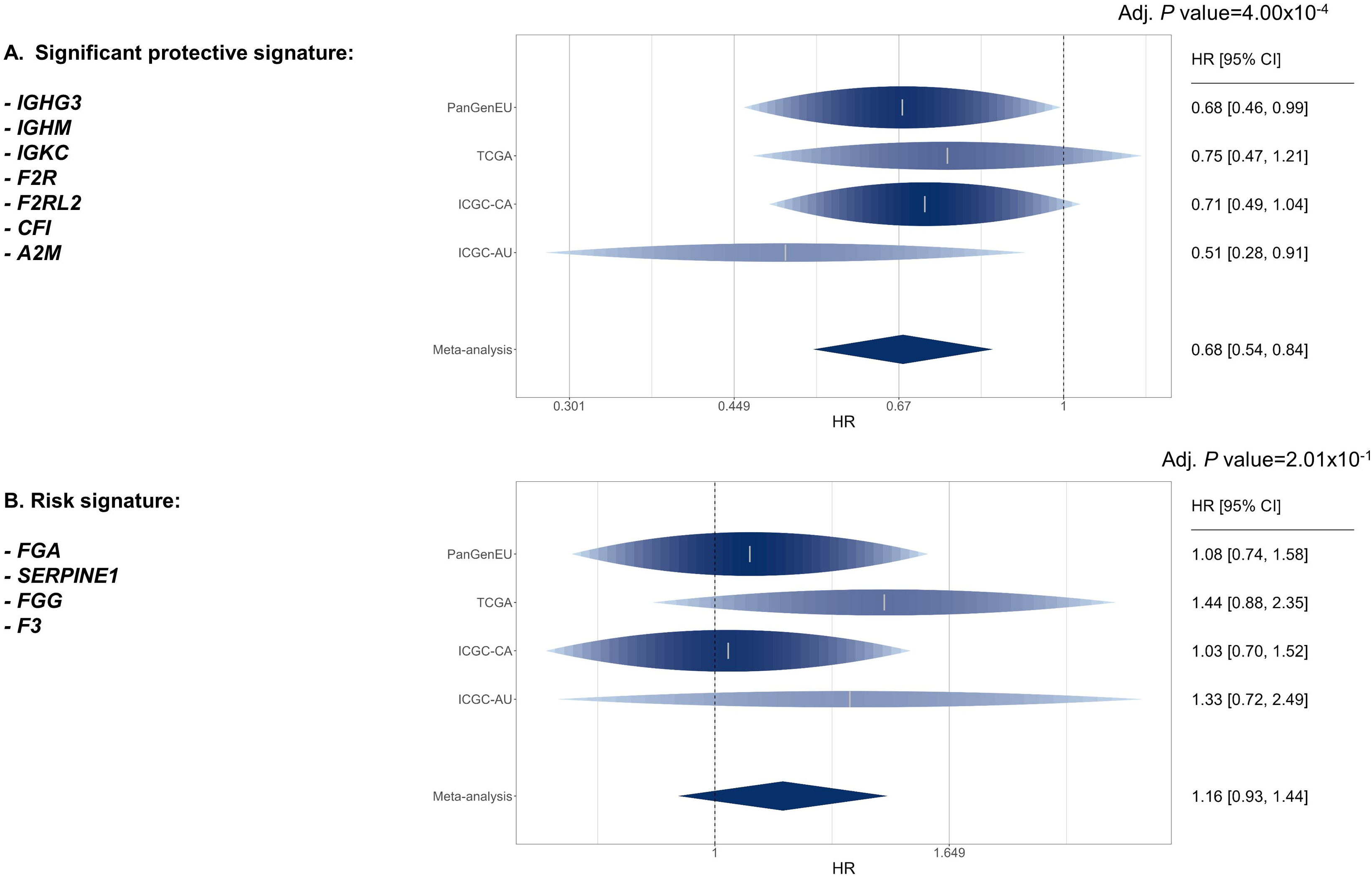
Forest plots from the meta-analysis of the CS-related genes for the protective signature comprising the CS-related genes associated with improved overall survival in the meta-analysis of PanGenEU, TCGA, ICGC-CA, and ICGC-AU (Fig. 3a), and the risk signature when considering CS-related genes associated with an increased risk of death in TCGA, ICGC-CA, and ICGC-AU cohorts (Fig. 3b). Hazard ratio (HR) and 95% confidence interval (CI) estimated with Cox proportional-hazards regression models under a dominance MoI.

### Patients stratified according to CS-related gene expression have distinct immune cell infiltration profiles

We found that patients with elevated expression of any of the CS-related genes associated with better survival in the meta-analysis (*IGHG3*, *IGKC*, *IGHM*, *F2R*, *F2RL2*, *CFI*, and *A2M*), had significantly higher infiltration of TCD8^+^, B cells, and Th1 (**Fig. 4**). These PDAC tumors also exhibited an increased abundance of cell states associated with improved survival like state-S1 TCD8^+^, state-S2 TCD4^+^, or state-S1 macrophages (**Fig. 5**), as well as a higher abundance of C10 ecotype (**Fig. 6**), also known to be associated with better survival.

**Fig. 4.**
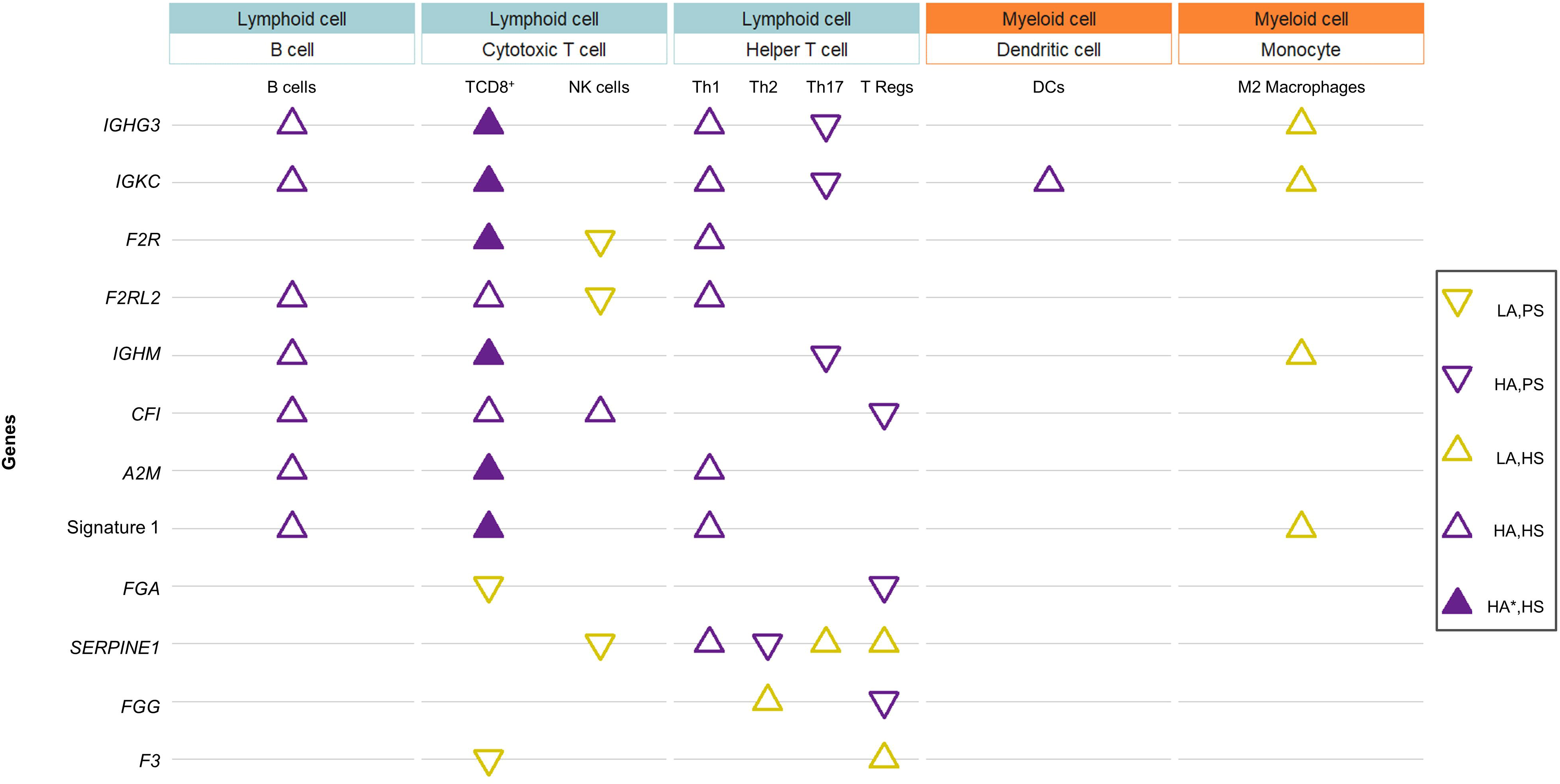
Bubble chart showing the significant differences in abundance of 9 immune cells when comparing high expression levels of the CS-related genes vs. low expression levels in any of the two study populations analyzed (TCGA and PanGenEU). LA (Lower Abundance) means that the immune cells are less infiltrated in the tumors with higher expression of the CS-related genes. HA (Higher Abundance) refers to a higher infiltration of the immune cells in PDAC tumors. PS (Shorter Survival) and HS (Longer Survival) corresponds to the expected survival based on the type of immune cell that is infiltrating the tumor and their abundance in the PDAC tumors. * The abundance of the infiltrated immune cells was significantly higher in both study populations.

**Fig. 5.**
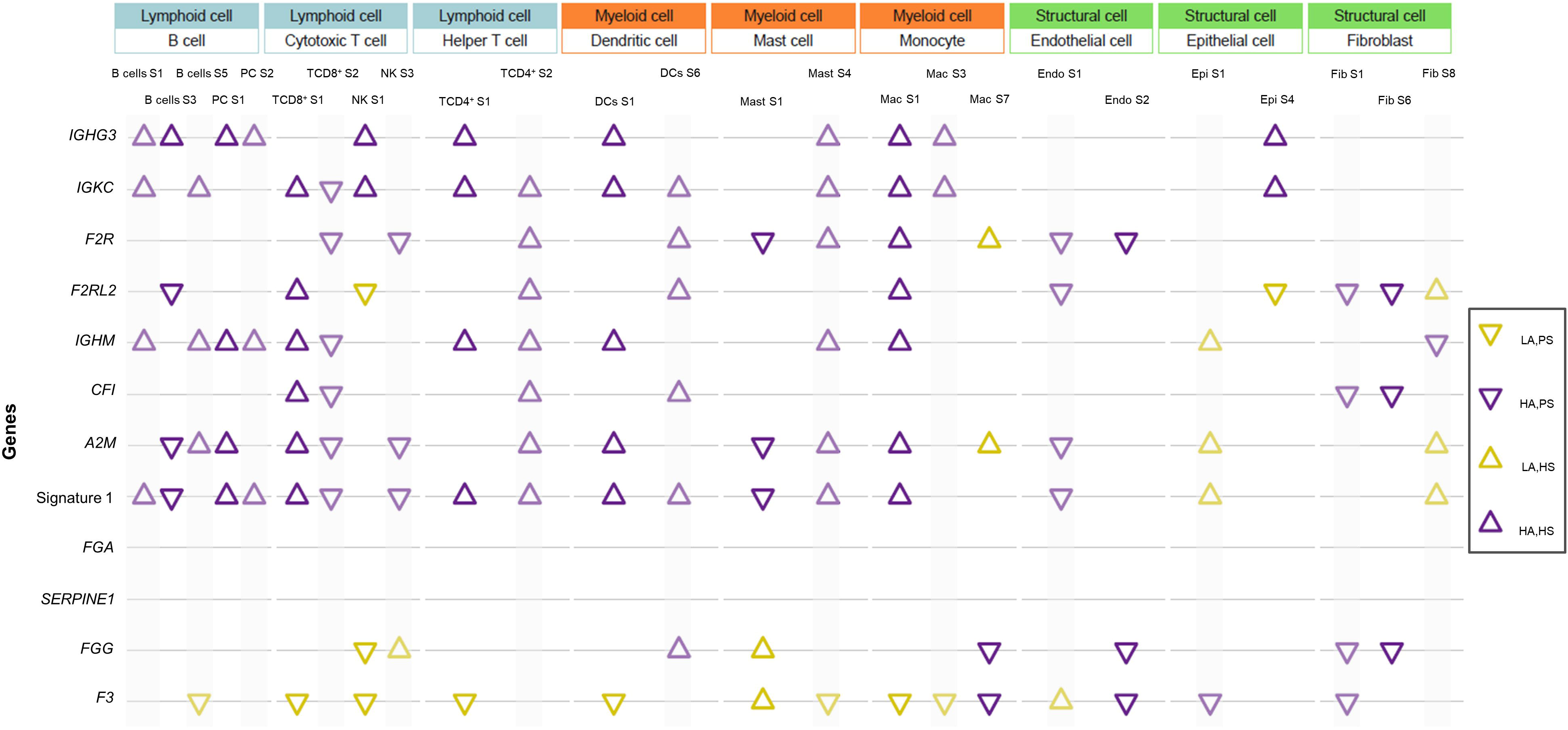
Bubble chart showing the significant differences in abundance of the cell states of 11 immune cells when comparing high expression levels of the CS-related genes vs low expression levels in the PanGenEU study. LA (Lower Abundance) means that the cell states are less prevalent in the tumors with higher expression of the CS-related genes. HA (Higher Abundance) refers to a higher prevalence of cell states in PDAC tumors. PS (Shorter survival) and HS (Longer survival) correspond to the expected survival based on the type of cell state for the different immune cells that are infiltrating the tumor and their abundance in the PDAC tumors.

**Fig. 6.**
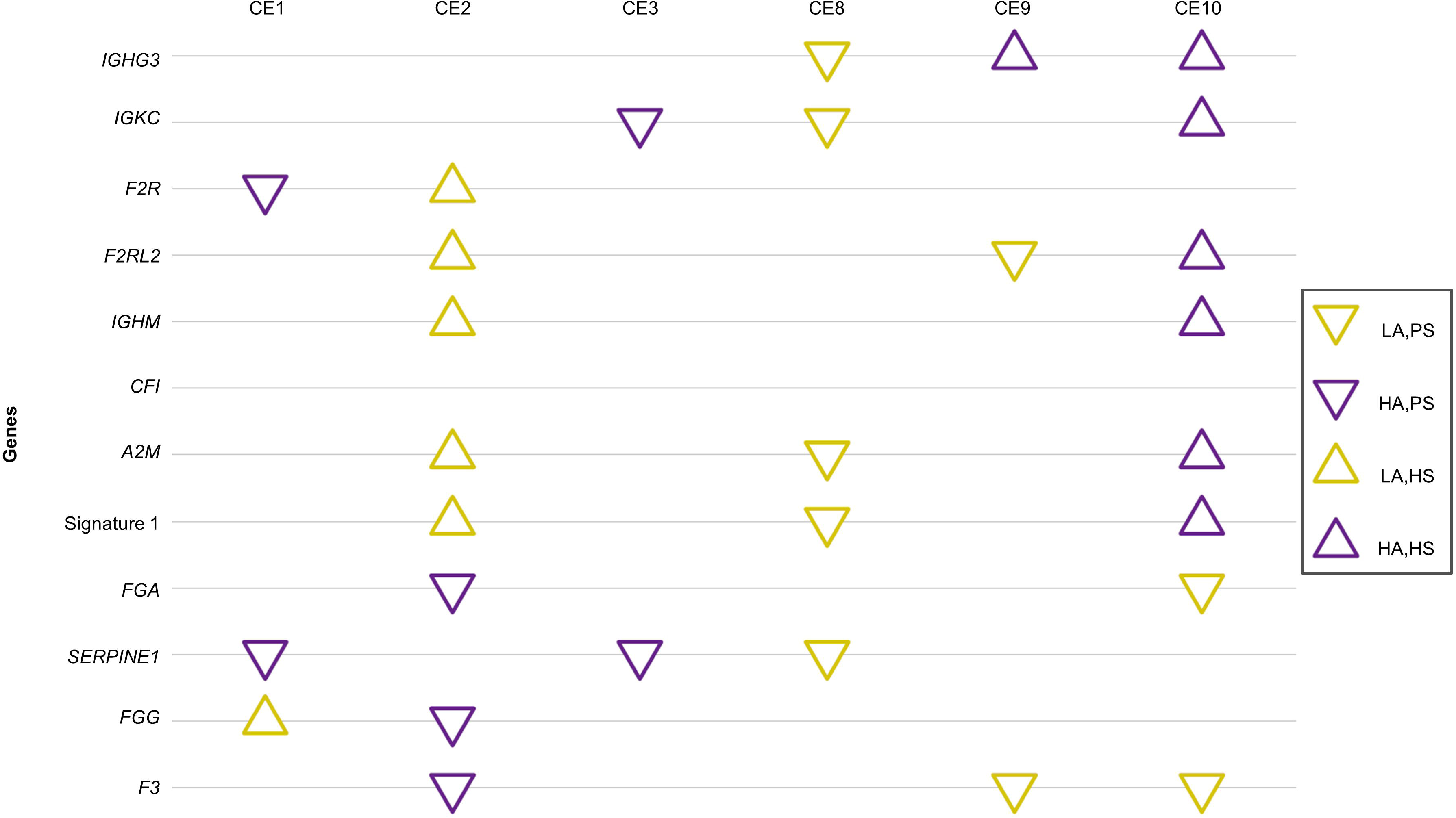
Bubble chart showing the significant differences in the abundance of the ecotypes (immune cell communities) when comparing high expression levels of the CS-related genes vs. low expression levels in the PanGenEU study. LA (Lower Abundance) means that the ecotypes are less prevalent in the tumors with higher expression of the CS-related genes. HA (Higher Abundance) refers to a higher prevalence of the ecotypes in PDAC tumors. PS (Shorter survival) and HS (Longer survival) correspond to the expected survival based on the ecotypes or cell communities that are infiltrating the tumor and their abundance in the PDAC tumors.

Patients with high expression of *Signature 1* also had increased infiltration by TCD8^+^, Th1, or B cells, and a reduced presence of cells like M2 macrophages (**Fig. 4**), higher abundance of state-S1 TCD8^+^, state-S1 and state-S2 TCD4^+^, and state-S1 B cells (**Fig. 5**), along with a higher abundance of ecotype CE10 (**Fig. 6**). Interestingly, PDAC tumors with high *Signature 1* levels showed elevated clonal expansion of B cells compared to those with low *Signature 1* levels (**Supplementary Fig. 4**).

Conversely, tumors expressing higher levels of *FGA* or *F3* genes, associated with worse outcomes, had significantly reduced infiltration of TCD8^+^ (**Fig. 4**). Tumors with high expression of *F3* or *FGG* had a lower abundance of cellular states and ecotypes associated with better survival outcomes, including state-S1 NK (**Fig. 5**) and CE10 ecotype (**Fig. 6**), and higher abundance of state-S1 mast cells and CE2 ecotype, associated with poorer survival.

## DISCUSSION

The development of pancreatic ductal adenocarcinoma is influenced by several genetic factors as described before^21,22^. The present study aimed to investigate the potential role of CS-related genes in PDAC risk and prognosis. Understanding the genetic factors involved in PDAC development is crucial to define high-risk populations suitable for screening and primary prevention targeted interventions.

Our findings provide valuable insights suggesting that CS is associated with PDAC risk as well as with of allergy and asthma through a meta-analysis of PanGenEU and UKBiobank study results, both at the SNP and gene levels. Functional *in-silico* analyses of identified susceptibility SNPs/genes have offered additional evidence supporting the involvement of CS-related genes in PDAC and shed light on their potential mechanisms of action, including immune regulation, expression modulation, and tumor-specific expression changes. In particular, eight SNPs in *VTN* identified as risk variants were also associated with a higher *VTN* expression in the normal pancreas. Our study also identified moderate and weak colocalization between PDAC and eQTL signals for *A2M* and *VTN*, respectively, suggesting that the SNPs within these genes might share a functional effect on PDAC risk and gene expression.

At the gene level, we identified six (four with borderline significance) susceptibility CS-related genes, including *FCN1*, which is expressed in leukocytes and involved in elastin-binding^23,24^; *PLAT* converts plasminogen into plasmin, crucial in cell migration and tissue remodeling/degradation^25^; *CD46* protects host-cell from complement self-cells damage^26,27^; *F2RL2* is key in thrombosis^28^; *VTN*, promotes cell adhesion^29^ and inhibits the MAC^30^; and *A2M*, which works as a protease inhibitor^31,32^. Remarkably, our study represents the first evidence of the association between these CS-related genes and PDAC risk. Interestingly, *PLAT* is among the top overexpressed genes in PDAC compared with normal pancreas^33,34^. Functional studies have shown its role in tumor growth and invasion in vitro and in vivo^35,36^ through a variety of mechanisms including non-catalytic effects^37^. We also discovered associations between *PLAT* and *F2RL2* and the risk of asthma, and *VTN* with the risk of allergy, highlighting the interplay between complement-related genetic variations and immune-related conditions also associated with PDAC risk.

Pathways related to humoral immune responses and coagulation are enriched with the identified CS-related genes. Notably, *A2M*, *F2RL2*, *VTN*, and *PLAT* are related to thrombosis, a condition that is prevalent in cancer patients, especially among those with PDAC^38^, who frequently suffer thromboembolic events^39–41^ due to the release of procoagulant factors like thrombin^42^. *A2M* plays a crucial role in regulating thrombin activity, while thrombin can cleave the *F2RL2* receptor^43^, enhancing platelet activation, promoting a procoagulant state, and contributing to thromboembolic diseases^44^. *VTN* binds to fibrin and platelets participating in platelet aggregation^45^ and inhibiting *PLAT*, responsible for dissolving blood clots^46,47^. The interplay between these genes and the coagulation cascade components warrants further investigation to elucidate their contribution to the development and progression of PDAC. Furthermore, DisGeNET analysis showed that some of these genes were associated with central nervous system diseases. *PLAT* and *A2M*, previously linked to depression^48,49^, were also associated with PDAC risk. Interestingly, PDAC has the highest prevalence of depression symptoms preceding its diagnosis^50–52^ compared to other cancer types, suggesting a potential connection between genetic variations in *PLAT* and *A2M*, depression, and PDAC. Furthermore, *A2M* has been linked to neuronal disorders like Alzheimer’s disease^53,54^. PDAC is a neurotropic tumor^55–57^ with a high incidence of perineural invasion. Given this neurotropic nature of PDAC, the association between *A2M* and PDAC becomes even more relevant for its potential involvement in Alzheimer’s disease, supporting the observed inverse relationship between neurodegenerative diseases and cancer^58–62^. However, further research is needed to explore the interplay between these genes, neuronal and mental diseases, and the development of PDAC.

Next, we aimed to identify robust prognostic biomarkers considering the CS-related genetic variation and transcriptomic data. Although no significant variants at the germline level were identified, our results found biomarkers at the somatic level. We observed that overexpression of eight CS-related genes was associated with a favorable prognosis. Among them, *IGHG3*, *IGKC*, and *IGHM* code for proteins essential for immunoglobulin (IG) integrity, indicating a robust immune response potentially through the clonal expansion of B cells and increased IG levels to target tumor cells. We also found that high expression levels of *F2R* and *F2RL2*, crucial for platelet activation and thrombotic responses, were also associated with anti-tumor immune cell infiltration including TCD8^+^, TCD4^+^, B cells, and DCs. We hypothesize that these inflammation processes, in the context of “cold” tumors like PDAC, might favor the activation and infiltration of the abovementioned TCD8^+^, DCs, and B cells, leading to improved patient survival. Conversely, Wu et al. (2024)^63^ identified *F2R* as a potential biomarker of adverse outcomes in gastric adenocarcinoma due to inflammatory processes. Elevated expression of *F2RL2* has been associated with improved prognosis in esophageal squamous cell carcinoma^64^, although with poorer survival in colorectal adenocarcinoma^65^ and gliomas^66^. Furthermore, we showed that *A2M* expression was associated with enhanced overall survival outcomes and anti-tumor immune cell infiltration (TCD8^+^, Th1, and B cells), consistent with findings in intrahepatic cholangiocarcinoma where its low expression was associated with unfavorable prognosis^67^. Lastly, we found that greater expression of *CFI* was associated with enhanced patient survival and infiltration of tumor tolerant immune cells (TCD8^+^, Th1, and NK cells). Other studies have associated *CFI* with shorter breast cancer survival^68^, and tumor progression in cutaneous squamous cell carcinoma and gliomas^69,70^. Considering that the dual role of these genes might be cancer-specific, further research on the roles of *F2R, F2RL2*, *A2M*, *CFI*, and *C4A* in PDAC prognosis is crucial. Additionally, we propose a prognostic gene expression signature composed of the genes associated with a favorable prognosis: *Signature 1*. Interestingly, patients with higher levels of this signature had tumors with enhanced clonal expansion, and tumor-killing immune cells were present, suggesting a more robust immune response. Based on these findings, we anticipate that these patients may respond favorably to immunotherapy.

We also identified four biomarkers associated with unfavorable prognosis: *FGA*, *FGG*, *SERPINE1*, and *F3*. These genes are involved in blood clot formation through the coagulation cascade. Higher levels of *FGA* and *FGG* are linked to poor prognosis in gastric cancer^71^. Serum levels of *FGG* are also suggested to act as a biomarker for castration-resistant prostate cancer^72^. *SERPINE1* expression has been associated with poorer prognosis^73^ in colon and cancer progression in colon^74^ and gastric^75^ cancer. *F3*, which activates the coagulation cascade via coagulation factor VII interaction, is associated with colorectal cancer^76^. We observed that PDAC tumors displaying high expression of these genes also had a decreased presence of anti-tumor immune cells like TCD8^+^ and cell communities such as CE1 and CE2^77^ ecotypes, possibly creating an environment conducive to tumor progression, leading to poorer patient outcomes. Interestingly, a signature comprising *SERPINE1* and *F3* among other genes, has been recently associated with worse prognosis in lower-grade glioma^78^. Based on our results and previous evidence, we speculate that targeting *SERPINE1*, *F3, FGG*, or *FGA* genes in combination with chemotherapies, could enhance treatment effectiveness in PDAC tumors. This is particularly relevant as some F3 targeted therapies Genmab’s tisotumab vedotin are already approved to treat cervical cancer^79^. Additionally, *SERPINE1* has been shown to reduce the efficacy of chemotherapy agents promoting drug resistance in triple-negative breast tumors^80^, while *FGA* has been proposed for targeted therapy responsiveness in NSCLC patients^81^.

This study has some limitations. First, our analyses focused on the Caucasian population, and therefore, the generalizability to other ancestry groups remains uncertain. Additionally, the study relied on self-reported epidemiological information, which may introduce misclassification of risk factors and comorbidity. However, the use of a meta-analysis approach incorporating data from different populations helped mitigate this potential bias. Another limitation is that we focused on common variants. Future studies should explore rare variants through exome or genome sequencing or targeted analysis to gain deeper insights. Lastly, the SKAT-O approach utilized in our study can determine the significance of the association between the genes and PDAC risk/prognosis, but it does not elucidate the direction of the effect. We minimized this limitation by applying functional *in silico* studies.

Importantly, our study has several strengths. We conducted an extensive analysis of the association between the CS-related genes and PDAC risk utilizing advanced methodologies in two large studies with distinct designs, PanGenEU and UK Biobank, enhancing the statistical power and robustness of our findings. Additionally, we thoroughly examined the association of CS-related genes with various PDAC risk factors using comprehensive epidemiological and clinical data, which provides a deeper understanding of the complex interplay between CS-related genes and PDAC risk. The use of four independent study populations to explore the role of CS-based genes on PDAC prognosis allowed us to identify a strong signature indicative of a better prognosis, which is also supported by immune infiltration characteristics within the tumor in addition to the statistical evidence.

In conclusion, our work exhaustively assesses the association of six CS-related genes (*CD46*, *A2M*, *VTN*, *F2RL2*, *FCN1*, and *PLAT*) with PDAC risk. The functional relevance of these genetic variations, particularly in gene expression regulation and disease pathways, underscores their role in PDAC. These findings highlight their importance as potential biomarkers for PDAC risk profiling, leading to early diagnosis. Furthermore, we showed that gene expression patterns of CS-related genes may help to discriminate PDAC patients based on their prognosis. We also shed light on the molecular mechanisms underlying pancreatic carcinogenesis, which might provide valuable insights for personalized treatment strategies. Future research should focus on unraveling the intricate mechanisms through which these complement-related genes contribute to PDAC development, as well as exploring their potential as therapeutic targets in PDAC tumors.

## Data Availability

All data produced in the present study are available upon reasonable request to the authors

## Acknowledgments

The authors are thankful to the patients, coordinators, field and administrative workers, and technicians of the European Study into Digestive Illnesses and Genetics (PanGenEU) study. We also thank Guillermo Pita and Anna González-Neira, from CEGEN-CNIO, and Joe Dennis and Laura Fachal, from the University of Cambridge, for genotyping PanGenEU samples, performing variant calling and SNP imputation, and editing data. This research has been conducted using the UK Biobank Resource under Application Number 47884. RR gratefully acknowledge funding from NIH (R35 CA253126.) and SU2C Convergence 3.14. This research has been conducted using the UK Biobank Resource under Application Number 47884. The results here are in part based upon data generated by the TCGA Research Network: https://www.cancer.gov/tcga.

## Authors contribution

Study conception: NM, ELM. Design of the work: AL, RR, ELM, NM. Data acquisition: IF, RTL, AC, RAG, MI, XM, JML, CM, JP, MO, VMB, AT, AF, LMB, TCJ, EFM, TG, WG, LS, JB, EC, JK, BK, JM, DO, AS, WY, FXR, NM, and rest of the PanGenEU Investigators. Data analysis: AL, LA, SS, AM, ELM. Interpretation of data: AL, RR, FXR, ELM, NM. Creation of new software used in the work: No one. Drafting the work or substantively revising it: AL, RR, FXR, ELM, NM. Approval of the submitted version (and any substantially modified version that involves the author’s contribution to the study): ALL AUTHORS. Agreement on both to be personally accountable for the author’s own contributions and to ensure that questions related to the accuracy or integrity of any part of the work, even ones in which the author was not personally involved, are appropriately investigated, resolved and the resolution documented in the literature: ALL AUTHORS.

## Availability of data and materials

The PanGenEU datasets analyzed during the current study are not publicly available due to the sensitive nature of genetic data but are available from the corresponding author upon reasonable request. Other datasets are UK Biobank, the International Cancer Genome Consortium (ICGC), Pancreatic Cancer Canada (ICGC-CA) and Pancreatic Cancer Australia (ICGC-AU) repositories (https://dcc.icgc.org/repositories), and The Cancer Genome Atlas (TCGA) repository (https://portal.gdc.cancer.gov).

