## Supplementary Material and Methods for "Deciphering the Role of Complement System Genes in Pancreatic Cancer Susceptibility and Prognosis"

**Additional File 1**

| **Supplementary Methods** | | **Page 3** |
| --- | --- | --- |
| **Annex 1.** | PanGenEU Investigators. | **Page 7** |
| *The following Supplementary Tables are provided in separate sheets of the excel file Supplementary tables.xlsx* | | |
| **Table S1.** | ICD9 and ICD10 codes for PDAC in UK Biobank. |  |
| **Table S2**. | List of 111 selected genes in the complement-related immunity gene-set. Adapted from Qian et al., 2019. |  |
| **Table S3.**  **Table S4.**  **Table S5.** | Baseline characteristics of the study populations.  SNP ID, position, gene, odds ratios (OR) and 95% confidence intervals bounds (ci.lb and ci.ub) of the single variant association meta-analysis between the CS-related SNPs and pancreatic ductal adenocarcinoma (PDAC) risk.  Number of SNPs (SNPs) within each gene and *P* value (*P*) of the association between the CS-related genes and known risk/protective factors for PDAC. |  |
| **Table S6.** | Posterior probabilities of the colocalization analysis of both traits PDAC and eQTL or sQTL. The posterior probabilities that meet the conditions for both traits are highlighted in bold font. |  |
| **Table S7.** | SNP ID, position, gene, hazard ratios (HR) and 95% confidence intervals bounds (ci.lb and ci.ub) of the single variant association meta-analysis between the CS-related SNPs and pancreatic ductal adenocarcinoma (PDAC) survival. |  |
| **Table S8.** | Table S8. Number of SNPs (nSNPs) within each gene and *P* value (*P*) of the SKAT-O association analysis between the CS-related genes and PDAC survival. |  |
| **Figure S1**. | Enrichment analysis of the susceptibility CS-related genes associated with PDAC through FUMA GWAS. | **Page 8** |
| **Figure S2**. | Gene-disease class heatmap showing the diseases previously found to be associated with our PDAC susceptibility CS-related genes. | **Page 8** |
| **Figure S3.** | eQTLs (Figure S3A) and sQTLs (Figure S3B) found in normal pancreatic cells within some of the CS-related genes significantly associated with PDAC risk in the meta-analysis. Those SNPs associated with an increase in the gene expression or with the predisposition to a specific isoform are shown in red, whereas those associated with a decrease in gene expression or a lower predisposition to a specific isoform are shown in blue. The SNPs that are both eQTLs/sQTLs and were included in our SKAT-O analysis are marked with a golden rhombus. | **Page 9** |
| **Figure S4.** | Differences in the expression levels of the CS-related genes between pancreatic tumour cells (purple) and pancreatic normal cells (orange). | **Page 9** |

**Supplementary methods**

**Study populations**

We utilized the resources of the PanGenEU case-control study, the UK Biobank, the International Cancer Genome Consortium (ICGC), Pancreatic Cancer Canada (ICGC-CA) and Pancreatic Cancer Australia (ICGC-AU) repositories (<https://dcc.icgc.org/repositories>), and The Cancer Genome Atlas (TCGA) repository (<https://portal.gdc.cancer.gov>).

*PanGenEU* study details have been reported elsewhere^1–3^. It is a multicenter case-control study on PDAC carried out in 28 centers across Spain, Italy, Sweden, Germany, the UK, and Ireland between 2009-2014 and 2016-2018, recruiting newly diagnosed PDAC patients aged 18 and older. Control subjects were sex-, age- and area of residence-matched hospital in-patients without cancer whose primary diagnoses were not associated with well-known PDAC risk factors. In Ireland and Sweden, population-based controls were recruited. Trained monitors conducted in-person interviews using standardized surveys to collect epidemiological information, including lifestyle factors, environmental exposures, and medical history.^1^ Participants with incomplete medical information were excluded from this analysis. DNA was extracted from buffy coats; genotyping was conducted at the CEGEN facility (Spanish National Cancer Centre, CNIO, Madrid, Spain) using the Infinium OncoArray-500K^4^. To impute the missing genotypes, IMPUTE2 v2^5^ with the 1000 Genomes Project Phase 3 (v1)^6^ as a reference was employed. Subjects with a high rate of missing calls, unexpected heterozygosity, discordance between reported and genotyped sex, unexpected relatedness, and estimated European ancestry below 80% were excluded from the study. This resulted in a final dataset comprising 1,317 cases and 700 controls for the association analysis with PDAC risk. Among PDAC cases, we had RNAseq data for 142 PDAC patients. RNA was extracted from manually microdissected FFPE tumor sections. The raw RNA sequencing data came from two distinct library preparations: the first dataset employed unique molecular identifiers for accurate mRNA quantification, while the second used the TruSeq RNA Access Library Prep Kit for paired-end sequencing. These approaches were complemented by variant calling from RNA-Seq data and whole exome sequencing to analyze genetic variations, contributing to a comprehensive genomic characterization of PDAC samples from the PanGenEU study. Finally, the reads were aligned with STAR^7^ to the GRCh38 GENCODE26 of the human genome. We performed a survival analysis including 122 PDAC patients who had complete clinical information. Additionally, data from 142 PDAC patients was employed to assess the immune landscape of these PDAC tumors.

*UK Biobank.* This is a prospective cohort study with genetic and phenotypic data from about 500,000 individuals from the United Kingdom^8^. Genotyping was performed using two arrays: the UK Biobank Axiom array for 450,000 subjects and the UK BiLEVE Axiom array for the remaining 50,000 individuals. The selection of cases was based on their primary codes according to the International Classification of Diseases (ICD-9 and ICD-10) (**Supplementary** **Table 1**). To ensure the reliability of PDAC diagnoses, self-reported PDAC cases (N=45) and individuals who did not have Caucasian genetic ancestry (N=129) were excluded, resulting in a final dataset of 761 PDAC cases. For the control group, individuals without a history of cancer and matching the same ICD9 and ICD10 codes as the PanGenEU controls were selected.^2^ Finally, we focused specifically on a subset of the population consisting of 95,811 individuals of Caucasian ancestry for the subsequent association analysis.

The *ICGC* is a worldwide consortium aiming at studying the genomics, of a wide variety of cancer types^9^. The ICGC-CA study consists of 160 patients across Canada with WGS and RNAseq data. ICGC-AU is composed of 86 PC patients with gene expression, clinical and mutational profiles obtained from ICGC data portal.

*TCGA* was established by the US National Institute of Health and is one of the world’s largest cancer genomics datasets. The Affymetix 6.0 SNP array was used to genotype 33 distinct cancer types. We collected information on SNPs within CS-related genes as well as clinical information from 134 PDAC cases. Additionally, immune populations of 134 TCGA PDAC patients were obtained from Thorsson et al, 2018^10^ and Pineda et al, 2021^11^ to assess the immune landscape of these tumors.

**In-silico functional analysis**

To understand the functional implications of the genes significantly associated with PDAC risk, we conducted an *in silico* functional analysis using various bioinformatics tools. First, we utilized FUMA GWAS^12^, a comprehensive tool for functional annotation and interpretation of GWAS results to analyze the pathways and biological processes enriched with the susceptibility genes identified in our gene-based association analyses. We employed DisGeNET^13^, a database that integrates information on gene-disease associations, to explore previously reported disease relationships with the identified genes. We also investigated the eQTL and sQTL within these genes in normal pancreas tissue from the Genotype-Tissue Expression (GTEx) database^14^. To assess whether the observed genetic signals were specific to PDAC risk or shared with gene expression/splicing regulation, we performed a colocalization analysis using the summary statistics of PDAC meta-analysis and eQTL-sQTL analysis in 243 normal pancreas from GTEx. We employed the function *coloc.abf* (*coloc* R package, v. 3.2-1)^15^.

**Survival analysis of CS-related genes at germline level**

We examined the association between the SNPs within 111 CS-related genes and PDAC overall survival (OS) in the PanGenEU and UK Biobank populations using a Cox proportional-hazards regression (Cox PH) model adjusted for sex, age at diagnosis and stage of the tumor. Then, we conducted a gene-based survival analysis using the SKAT-O model on SNPs from the single variant association analysis, also adjusted for the aforementioned covariates. Results from both analyses were then meta-analyzed using a random-effects model across the two populations.

**Immune infiltration in PDAC tumors**

We explored whether the observed associations between CS-related genes at the individual and signature levels with survival might be partially explained by specific immune infiltration patterns within PDAC tumors. We employed TCGA (N=134) data from Thorsson et al, 2018 and Pineda et al, 2021 and utilized CIBERSORTx^16^ in the PanGenEU population (N=142) to assess the abundance of tumor infiltrating immune cells. Next, the ECOTYPER^17^ was applied in the PanGenEU population to discern the cellular states of the infiltrating cells and study the immune communities known as ecotypes from gene expression data. Patients were stratified based on the expression levels of *IGHG3*, *IGHM*, *IGKC*, *F2R*, *F2RL2*, *CFI*, *A2M*, *FGA*, *SERPINE1*, *FGG*, and *F3* (high vs low for each gene). We employed the Wilcoxon test to examine differences in the immune landscape (infiltrating immune cells, cell states, and ecotypes) between the two groups. Significant differences were defined by a *P* < 0.05. The same approach was applied to evaluate differences in immune cell infiltration between patients scoring low-expression vs high-expression on the prognostic signature.

**Annex 1. PanGenEU Centres and Investigators**

Spanish National Cancer Research Centre (CNIO), Madrid, Spain: Núria Malats^1^, Francisco X Real^1^, Evangelina López de Maturana, Paulina Gómez-Rubio, Esther Molina-Montes, Lola Alonso, Mirari Márquez, Roger Milne, Ana Alfaro, Tania Lobato, Lidia Estudillo.

Verona University, Italy: Rita Lawlor^1^, Aldo Scarpa, Stefania Beghelli.

National Cancer Registry Ireland, Cork, Ireland: Linda Sharp^1^, Damian O’Driscoll.

Hospital Madrid-Norte-Sanchinarro, Madrid, Spain: Rafael, Álvarez^1^, Manuel Hidalgo, Jesús Rodríguez Pascual.

Hospital Ramon y Cajal, Madrid, Spain: Alfredo Carrato^1^, Carmen Guillén-Ponce, Mercedes Rodríguez-Garrote, Federico Longo-Muñoz, Reyes Ferreiro, Vanessa Pachón, M Ángeles Vaz.

Hospital del Mar, Barcelona, Spain: Lucas Ilzarbe^1^, Cristina Álvarez-Urturi, Xavier Bessa, Felipe Bory, Lucía Márquez, Ignasi Poves🕇, Fernando Burdío, Luis Grande, Mar Iglesias, Javier Gimeno.

Hospital Vall d´Hebron, Barcelona, Spain: Xavier Molero^1^, Luisa Guarner🕇, Joaquin Balcells.

Technical University of Munich, Germany: Christoph Michalski^1^, Jörg Kleeff, Bo Kong.

Karolinska Institute, Stockholm, Sweden: Matthias Löhr^1^, Jiaqui Huang, Weimin Ye, Jingru Yu.

Hospital 12 de Octubre, Madrid, Spain: José Perea^1^, Pablo Peláez.

Hospital de la Santa Creu i Sant Pau, Barcelona, Spain: Antoni Farré^1^, Josefina Mora, Marta Martín, Vicenç Artigas, Carlos Guarner, Francesc J Sancho, Mar Concepción, Teresa Ramón y Cajal.

The Royal Liverpool University Hospital, UK: William Greenhalf^1^, Eithne Costello.

Queen’s University Belfast, UK: Michael O’Rorke^1^, Liam Murray🕇, Marie Cantwell.

Laboratorio de Genética Molecular, Hospital General Universitario de Elche, Spain: Víctor M Barberá^1^, Javier Gallego.

Instituto Universitario de Oncología del Principado de Asturias, Oviedo, Spain: Adonina Tardón^1^, Luis Barneo.

Hospital Clínico Universitario de Santiago de Compostela, Spain: Enrique Domínguez Muñoz^1^, Antonio Lozano, Maria Luaces.

Hospital Clínico Universitario de Salamanca, Spain: Luís Muñoz-Bellvís^1^, J.M. Sayagués Manzano, M.L. Gutíerrrez Troncoso, A. Orfao de Matos.

University of Marburg, Department of Gastroenterology, Phillips University of Marburg, Germany: Thomas Gress^1^, Malte Buchholz, Albrecht Neesse.

Queen Mary University of London, UK: Tatjana Crnogorac-Jurcevic^1^, Hemant M Kocher, Satyajit Bhattacharya, Ajit T Abraham, Darren Ennis, Thomas Dowe, Tomasz Radon

Scientific advisors of the PanGenEU Study: Debra T Silverman (NCI, USA) and Douglas Easton (U. of Cambridge, UK)

^1^ Principal Investigator in each centre

**Figure S1.** Enrichment analysis of the susceptibility CS-related genes associated with PDAC through FUMA GWAS.

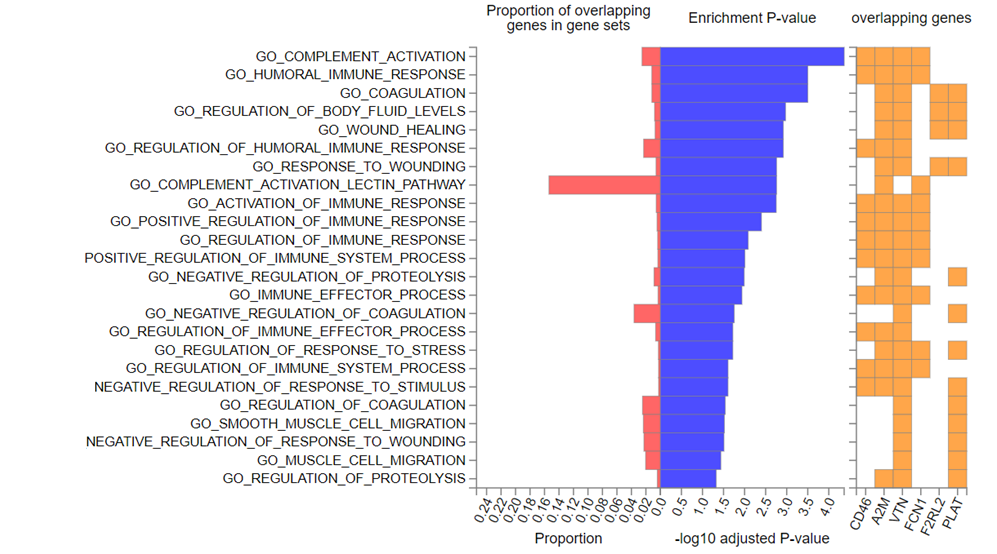

**Figure S2.** Gene-disease class heatmap showing the diseases previously found to be associated with our PDAC susceptibility CS-related genes.

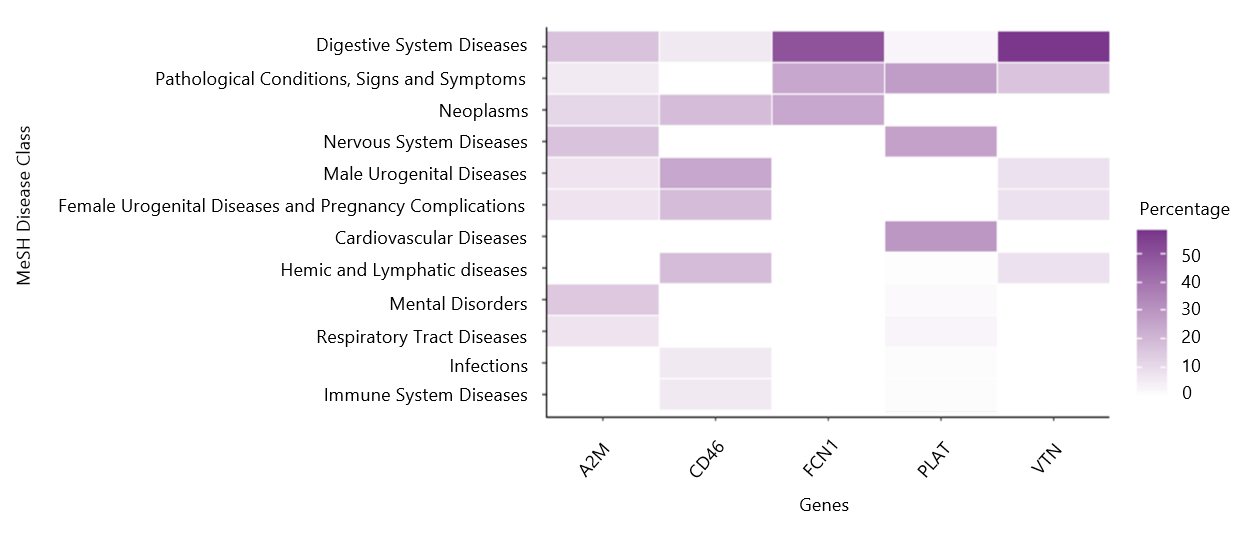

**Figure S3.** eQTLs (Figure S3A) and sQTLs (Figure S3B) found in normal pancreatic cells within some of the CS-related genes significantly associated with PDAC risk in the meta-analysis. Those SNPs associated with an increase in the gene expression or with the predisposition to a specific isoform are shown in red, whereas those associated with a decrease in gene expression or a lower predisposition to a specific isoform are shown in blue. The SNPs that are both eQTLs/sQTLs and were included in our SKAT-O analysis are marked with a golden rhombus.

**A)**

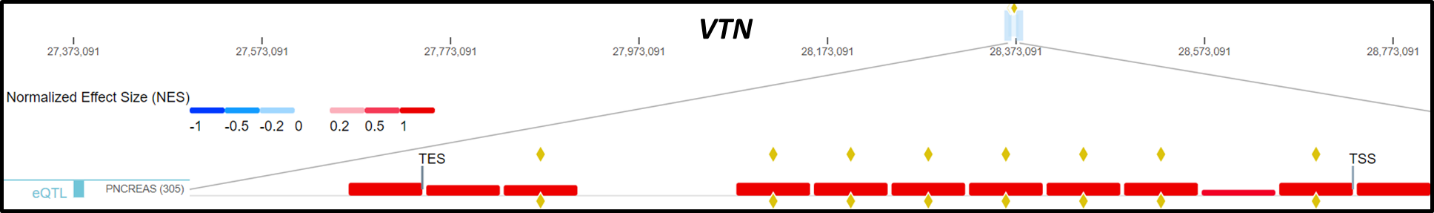

**B)**

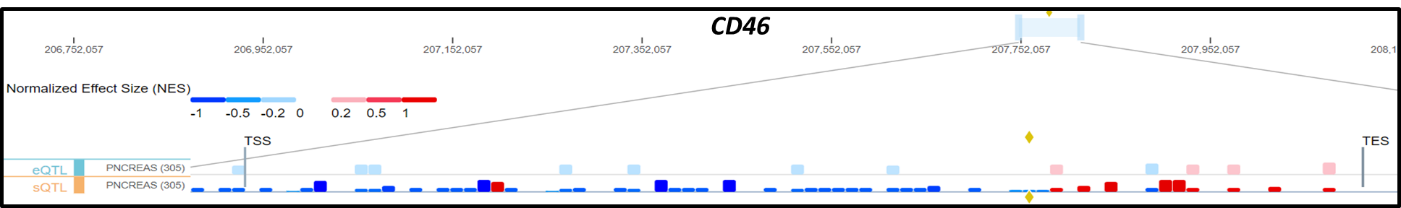

**Figure S4.** Boxplot showing the significant differences in clonal expansion levels between PDAC patients with high levels of the favourable *signature 1* vs low signature levels obtained through the Wilcoxon test.

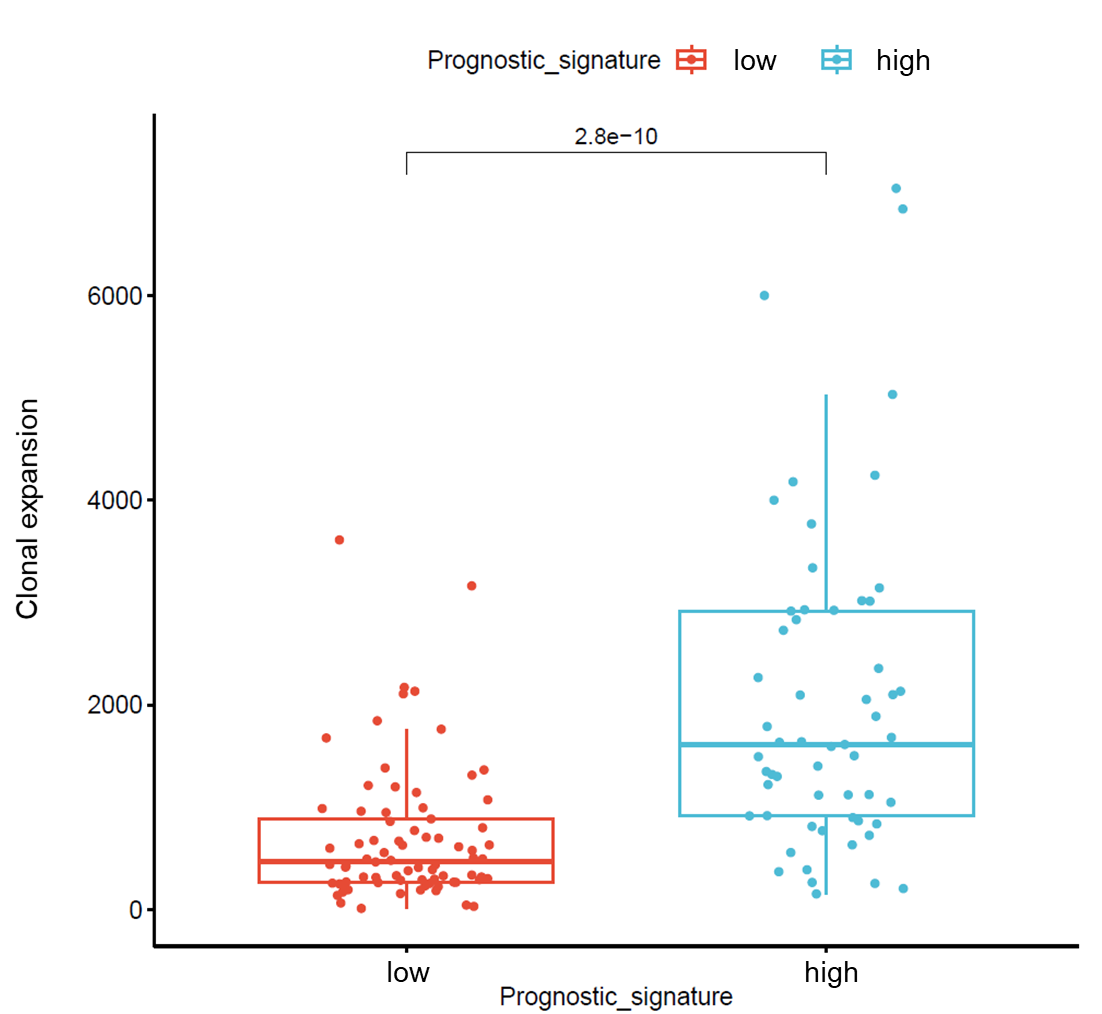
